# Clinical Characteristics and Risk factors for developed COVID-19 patients transferring to designated hospital from Jianghan Fangcang shelter Hospital: a retrospective, observational study

**DOI:** 10.1101/2020.04.21.20074724

**Authors:** Yunfei Liao, Yong Feng, Bo Wang, Hanyu Wang, Jinsha Huang, Yaxin Wu, Ziling Wu, Xiao Chen, Chao Yang, Xinqiao Fu, Hui Sun

## Abstract

**Background:** The outbreak of coronavirus disease 2019 (COVID-19) has become a world-wide emergency. Fangcang shelter hospitals have been applied in COVID-19 to ease ongoing shortage of medical resources in Wuhan since February 2020.

**Method:** This study enrolled all cases (no=1848) with mild or moderate type of COVID-19 in Fangcang shelter hospital of Jianghan in Wuhan from Feb 5th to Mar 9th, 2020. Diagnosis of COVID-19 was based on the National health commission of China. Epidemiological history, comorbidity, vital signs, symptoms and signs were recorded in detail. Laboratory tests included biochemical indicators and nucleic acid tests by throat swabs have been performed as well.

**Finding:** A total of 1327 patients reached the criteria of isolation release. Meanwhile, 521 patients have been transferred to the designated hospitals for further treatment, including severe type, fever more than 3 days, and severe comorbidity. The case-severity rate (rate of mild or moderate type transforming to severe type) was 3.0% in the shelter hospital. The patients from mild or moderate type to severe type showed the following clinical characteristics: the median incubation (onset to shelter) period was 10 days; they were all symptomatic at admission; fever, cough, and fatigue were the most common symptoms; hypertension, diabetes and coronary heart diseases were common co-morbidities; most of the patients had elevated levels of CRP at ill onset with 33.3% over 10 mg per L; bilateral distribution and ground-glass opacity were the most common manifestations in chest CT.

**Interpretation:** The potential risk factors of fever, fatigue, high level of C-reactive protein were the risk factors to identify the progression of COVID-19 patients with mild or moderate type. Fangcang shelter hospitals have substantially reduced the time from the onset of severe symptoms transfer to a designated hospital. Early application of the Fangcang shelter hospital may contribute to decrease the ratio of mild transforming to severe patients.

**Funding:** No specific grant from any funding was applied to this research.

**Research in context:** *Evidence before this study:* We searched PubMed from Nov 1, 2019, to Apr 8, 2020, for studies published in any language using the terms “COVID-19”, “coronavirus disease 2019”, “novel coronavirus”, “cabin hospital”, “shelter hospital”. Five studies have been found about coronavirus disease 2019 (COVID-19) in shelter hospital or cabin hospital. Fangcang shelter hospital of Jianghan received the largest number of patients among Fangcang shelter hospitals in Wuhan. These studies were related to development of Fangcang shelter hospitals, explaining three key characteristics (rapid construction, massive scale, and low cost) and five essential functions (isolation, triage, basic medical care, frequent monitoring and rapid referral, and essential living and social engagement). To our knowledge, there are no studies to comprehensively investigate a cohort of mild COVID-19 patients transfer to designated hospital from shelter hospital and their distinctive clinical features. Since Fangcang shelter hospital is a novel public health strategy, we aimed to investigate the clinical characteristics and risk factors for developed COVID-19 patients transfer to the designated hospital in Jianghan Fangcang shelter Hospital.

*Added value of this study:* From Feb 5th to Mar 9th, a total of 1848 cases of mild or moderate type of COVID-19 were enrolled in Fangcang shelter hospital of Jianghan (Wuhan, China). Of these cases, 521 patients were transferred to designated hospitals. Rate of mild or moderate type transforming to severe type was 3.0 % (56/1848) in the Fangcang shelter hospital. The median incubation (onset to shelter) period was 10 days (IQR 8.0-16.0). Patients with fever on cabin admission, high level C-reactive protein were also associated with mild-to-severe. Early application of the shelter hospital may contribute to alleviate the shortage of medical resources and decrease the ratio of severe patients. Furthermore, Fangcang shelter hospitals are likely to have substantially reduced the time from the onset of severe symptoms to admission to a designated hospital. The clinical characteristics of patients transferred to the designated hospital were important for the revision of admission criteria of COVID patients in Fangcang shelter hospitals. Dynamic observation the risk factors of mild to severe patients is contribute to great value for early prognosis and treatment.

*Implications of all the available evidence:* Keep vigilance of those mild patients whose had a fever over 38.0°C, cough and fatigue when they isolated at home. Fangcang shelter hospital could provide the rational strategy for isolation and triage of infected patients and decrease the family or community transmission cases.

## Introduction

The coronavirus disease 2019 (COVID-19) has been shown the ability of human-to-human transmission and become a world-wide emergency.^1^ The World Health Organization (WHO) has recently declared COVID-19 outbreak in several countries. Since January 2020, thousands of new patients have been diagnosed every day, which requires enormous medical resources. The surge of infections placed huge pressure on the national medical system.^2^

The Fangcang shelter hospitals in Wuhan were large-scale, temporary hospitals, rapidly built by converting existing public venues, such as exhibition centers and stadiums, into health-care facilities. They were served to isolate patients with mild or moderate COVID-19 from their families and communities, while providing basic medical care, disease monitoring, food, shelter, and social activities.^3^ Fangcang shelter hospitals presented five essential functions: isolation, triage, basic medical care, frequent monitoring and rapid transfer, and essential living and social engagement. Patients with mild or moderate COVID-19 who met additional admission criteria were isolated and treated in the Fangcang shelter hospitals, whereas patients with severe or critical COVID-19 received medical care in traditional hospitals.^3-6^ Fangcang shelter hospitals provide basic medical care and monitored the progression of disease. As some patients remain experienced progression of COVID-19 or development of severe chronic diseases, they were transferred in a timely manner to the designated higher-level hospitals. The clinical characteristics of patients transferred to the designated hospital were important for the revision of admission criteria of COVID patients in Fangcang shelter hospitals.

The case-severity (from mild or moderate to severe case) rate was an important benefit index for therapeutic efficacy assessment in shelter hospital. Dynamic observation the risk factors of mild to severe patients is contribute to great value for early prognosis and treatment. Therefore, a retrospective review of overall medical record was performed in Fangcang shelter hospital of Jianghan, which received the largest number of patients among Fangcang shelter hospitals in Wuhan. A total of 1848 cases with mild or moderate COVID-19 were included and 521 cases transferred to the designated hospital were analyzed. The risk factors of patients from mild or moderate to severe case were detected as well.

## Methods

### Patient selection

The Jianghan Fangcang shelter hospital opened on the 5^th^ Feb and closed on the 9^th^ Mar 2020. A total of 1848 cases with COVID-19 were enrolled in Jianghan Fangcang shelter hospital of Wuhan from Feb 5th to Mar 9th, 2020. The admission criteria of Fangcang shelter hospital were COVID-19 patients with mild or moderate type. Diagnosis of COVID-19 was based on the National health commission (NHC) of the People’s Republic of China.^7^ The clinical classifications are as follows: (1) mild, the clinical symptoms are mild and no pneumonia manifestation can be found in imaging; (2) moderate, patients have symptoms like fever and respiratory tract symptoms, etc. and pneumonia manifestation can be seen in imaging; (3) severe, meet any of the following conditions: a) respiratory distress, respiratory rate≥30 breaths / min; b) the oxygen saturation ≤ 93% at a rest state; c) arterial partial pressure of oxygen/ oxygen concentration (FiO2) ≤ 300mmHg (1mmHg = 0.133kPa); d) pulmonary imaging with >50% lesions progression within 24 to 48 hours. The release of isolation and discharge standards are as follows: (1) with normal body temperature for more than 3 days; (2) with significantly recovered respiratory symptoms; (3) lung imaging shows obvious absorption and recovery of acute exudative lesion; (4) with negative results of the nucleic acid tests of respiratory pathogens for consecutive two times (sampling interval at least 1 day).

This study was approved by the Research Ethics Commission of Wuhan Union Hospital ([2020]0038) and informed consent was waived by the Ethics Commission of Wuhan Union Hospital for emerging infectious diseases.

### Baseline data collection

Before admission into Fangcang shelter hospital, all suspected patients of Covid-2019 were taken upper respiratory throat swab samples. Chest CT scan was performed as well. Clinical and laboratory findings were recorded and carefully checked. Laboratory tests included biochemical Indicators, blood routine, and C-reactive protein (CRP, normal range 0-4 mg per L). Epidemiological history, comorbidity, vital signs, symptoms and signs were recorded in detail. Patients meeting the diagnosis of mild or moderate type were admitted to the mobile cabin hospital. Two reviewers (HW, JH) independently checked all collected data. In case of disagreement among two reviewers, consensus was conducted by a third reviewer (YW).

### Follow-up

During the following days in Fangcang shelter hospital, the patients were re-examined for laboratory and imaging examination, and recorded symptoms, signs, treatments and outcome events. The throat swab specimens of RT-PCR test and chest CT scan were performed according to the symptoms and signs. The clinical outcomes of patients in the mobile cabin hospital were divided into three ways. They were the patients transferred to the designated hospitals for further treatments, the patients reaching the criteria of isolation release, and the patients kept treatment in the mobile cabin.

### COVID-19 nucleic acid detection and chest CT scan

Throat swab samples were stored in virus transport medium and transported to Wuhan Union Hospital for laboratory diagnosis. Throat swab specimens of all patients were subject to real time PCR tests by amplifying ORF1ab gene and N gene of SARS-CoV-2 (BioGerm, Shanghai, China). The CT examinations were carried out with a 16-row multidetector CT scanner (μCT550, Shanghai LianYing medical technology co., LTD) using the following parameters: detector collimation widths 64 ×0.6 mm, 128×0.6 mm, 64×0.6 mm, and 64×0.6 mm; and tube voltage 120 kV.

### Statistical analysis

Continuous variables were represented by mean (standard deviations, SDs) or median (interquartile range, IQR) as appropriate, categorical variables were described as number (%). Significant differences between the 2 groups (mild patients and mild to severe patients) were compared by Student *t* test, Mann-Whitney U test, chi-square or Fischer exact test where appropriate. In addition, we also used univariable and multivariable logistic regression models to explore potential risk factors related to mild developing to severe. Considering the total included severe patients (n=56) in this study and in order to avoid over-fitting in our multivariable logistic regression models, we chose five variables for this model based on significant results (*P*<0.05) from the univariate regression. The results were present as odds ratio (OR) [95% confidence interval]. Because all tests were two-sided, *P* value less than 0.05 was considered statistically significant. Analyses were performed using SPSS 20.0 statistical package.

## Results

### General features of Jianghan Fangcang shelter hospital

According to the spatial structure, Jianghan Fangcang shelter hospital was is divided into 4 districts. Each district contains about 30 rooms and each room had 10-16 beds. About 300-400 patients were kept in each district per day. At least one nurse was assigned to each room and the distance between the neighboring beds was 1.5-1.8 meters in the room. There were about 25 doctors and 120 nurses on duty by turns in each distract, ensuring that doctors and nurses were available 24 hours a day. Doctors and nurses carried out the daily round separately. The digital ratio of patients to doctors and nurses was about 1.6 (1848/1180). After admission, all patients were given antiviral therapy (e.g., abidor hydrochloride) and other individualized treatments (such as antibiotics, antihypertensive and hypoglycemic therapy) according to the doctor’s advice and NHC’s interim guidelines.^7^

### Outcomes and Case-severity rate of patients in Fangcang shelter hospital

Among 1848 enrolled patients, the age range was from 15 to 81 years, and 49.0% were men. From Feb 5th to Mar 9th, 521 patients were transferred to designated hospitals for further treatment. Meanwhile, the other 1327 patients reached the criteria of isolation release or discharge, (Figure 1). Among 521 patients transferring to the designated hospitals, 10.7% patients with severe type from mild or moderate type (56 cases), 6.9% (36 cases) patients with body temperature more than 38.5°C for 3 days or more after treatment, 15.0% (78 cases) patients with cancer or severe liver/kidney/heart disease, 45.9% (239 cases) patients with the persistent positive nucleic acid testing after 2 weeks treatment, and 21.5% (112 cases) patients with other reasons (including new onset severe symptoms or mental illness or tremendous mental pressure or pregnant woman). The basic clinical characteristics of patients transferring to the designated hospitals were in Table 1.

**Table 1:**
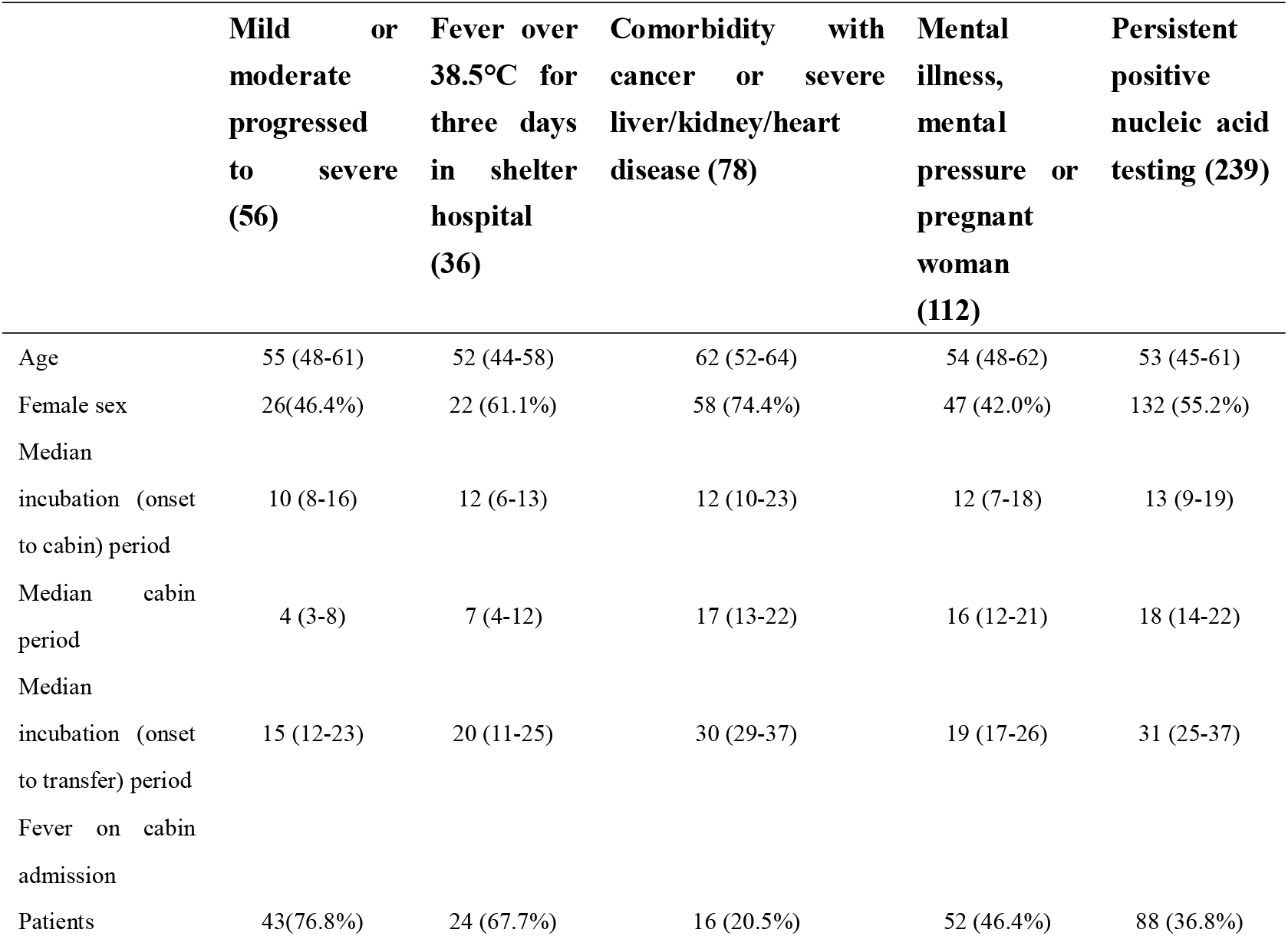
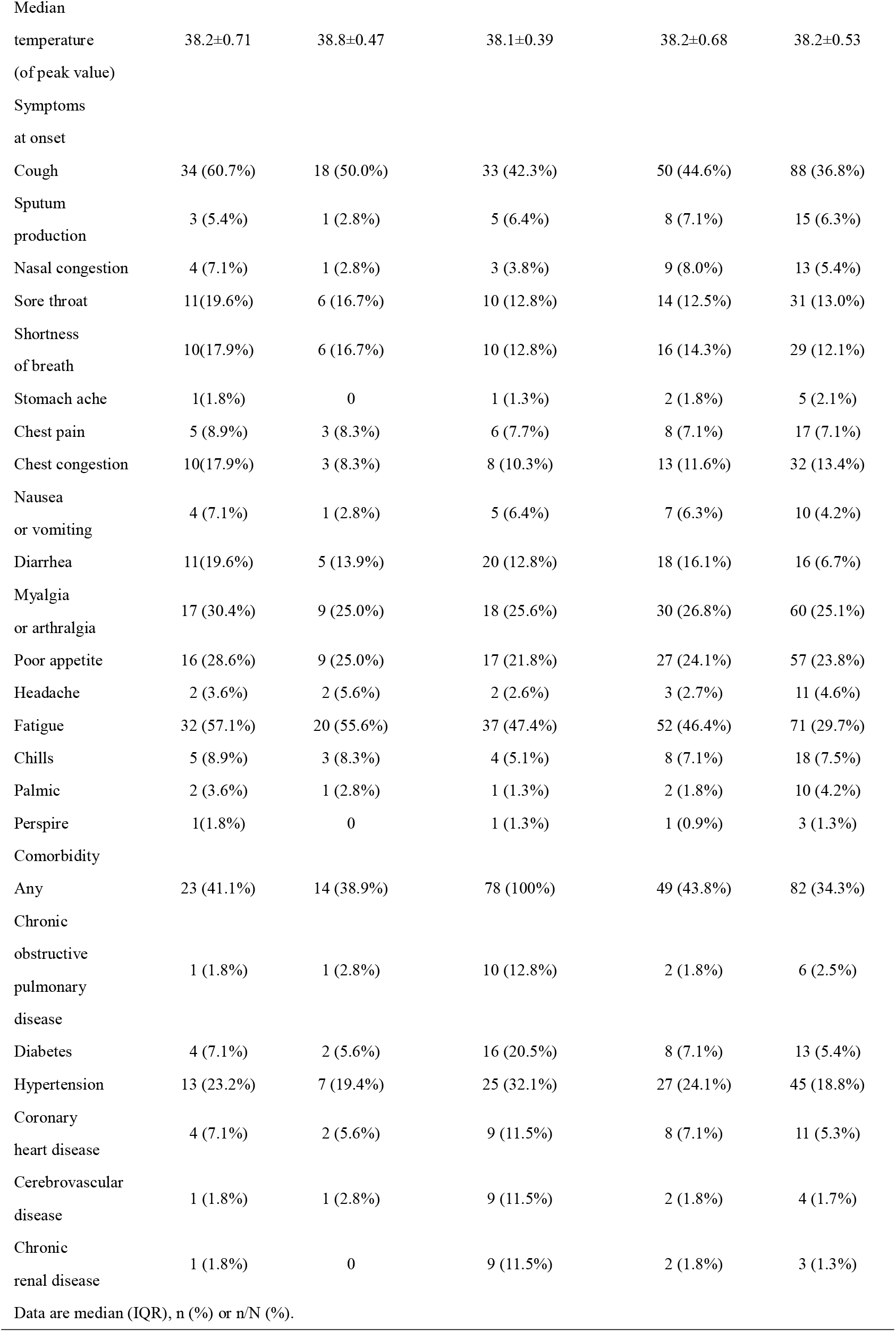
Clinical Characteristics of Patients Transferring to the Designated Hospitals from Fangcang Shelter Hospital.

**Figure 1:**
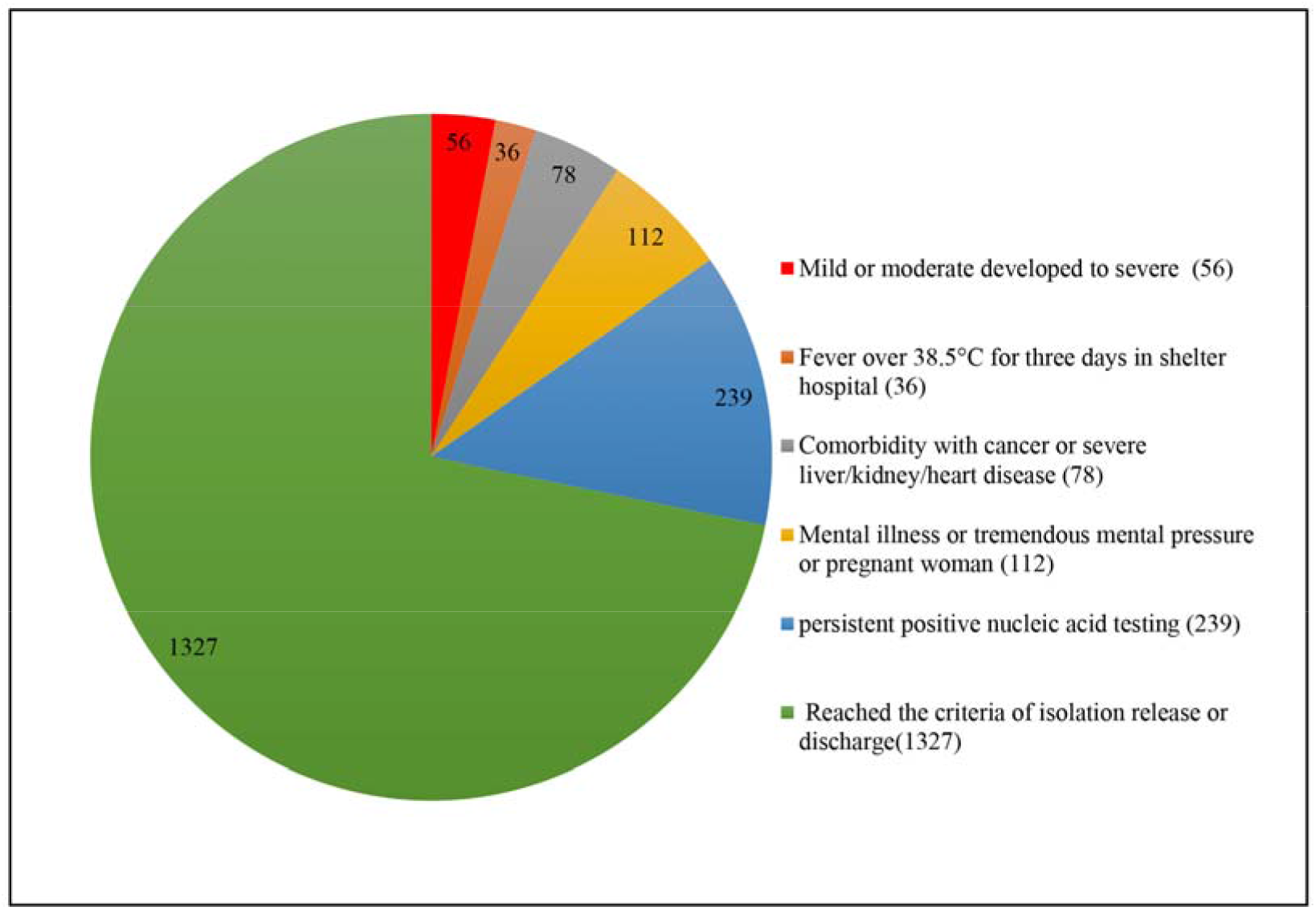
Outcome and distribution of all enrolled patients in Jianghan Fangcang shelter hospital (1848)

The case-severity rate was identified as the proportion of mild or moderate type progressing to severe type in this study. A total of 56 patients have progressed from mild or moderate type to the severe type. The case-severity rate of COVID-19 was 3.0% (56/1848) in the Fangcang shelter hospital of Jianghan.

### Clinical characteristics of patients transforming from mild or moderate type to severe type

In order to better display the clinical characteristics of mild or moderate to severe patients, 92 age-and sex-matched mild or moderate patients stayed in the Fangcang shelter hospital at the same time have been showed and compared as well. The severe patients have the following characteristics. Among 56 severe patients, 30 were male (53.6%) and 26 were female (46.4%). All the patients aged 28-73 years with an average age of 55 years (IQR 48.0-61.0). About 76.8% of those patients was between 40 to 65 years. The median incubation time (onset to severe) was 10 days (IQR 8.0-16.0), 7/56 (12.5%) less than 7 days, 34/56 (60.7%) between 7-14 days, 14/56 (25.0%) more than 14 days. Meanwhile, the median time in mobile cabin was 4 days (IQR 3.0-8.0), 12/56 (21.8%) less than 3 days, 29/56 (52.7%) between 3-7 days, 14/56 (25.5%) more than 7 days. (Table 2)

**Table 2:** Clinical Characteristics of the Mild or Moderate Progressed to Severe Patients.

|  | Mild or moderate progressed to severe (56) | Match mild or moderate (92) | *P* value |
| --- | --- | --- | --- |
| Age |  |  |  |
| Median (IQR) — year | 55 (48-61) | 56 (48-62) | 0.909 |
| Distribution of age— no./total no. (%) | .. | .. | 0.976 |
| <40 | 7 (12.5%) | 11 (12.0%) | .. |
| 40-65 | 43 (76.8%) | 72 (78.3%) | .. |
| >65 | 6 (10.7%) | 9 (9.8%) | .. |
| Female sex — no./total no. (%) | 26(46.4%) | 48 (52.2%) | 0.498 |
| Smoking history — no./total no. (%) | 3 (5.4%) | 5 (5.4%) | 0.984 |
| Median incubation (onset to shelter) period (IQR) — days | 10 (8-16) | 14 (11-19) | <0.0001 |
| Distribution of period— no./total no. (%) | .. | .. | 0.016* |
| <3days | 7(12.5%) | 9 (9.8%) | .. |
| 3-7days | 34 (60.7%) | 36 (39.1%) | .. |
| >7days | 14 (25.0%) | 44 (47.8%) | .. |
| Median cabin period (IQR) — days | 4 (3-8) | 17 (12-22) | <0.0001 |
| Distribution of period — no./total no. (%) | .. | .. | <0.0001* |
| <3days | 12 (21.8%) | 1 (1.1%) | .. |
| 3-7days | 29(52.7%) | 10 (11.0%) | .. |
| >7days | 14 (25.5%) | 80 (87.9%) | .. |
| Median incubation (onset to transfer) period (IQR) — days | 15 (12-23) | 33 (28.5-37) | <0.0001 |
| Distribution of period — no./total no. (%) | .. | .. | <0.0001* |
| <7days | 2 (3.6%) | 0 | .. |
| 7-14days | 24 (43.6%) | 2 (2.2%) | .. |
| >14days | 29 (52.7%) | 87 (97.8%) | .. |
| Fever on cabin admission |  |  |  |
| Patients — no./total no. (%) | 43(76.8%) | 24 (26.1%) | <0.0001 |
| Mean temperature (of peak value) (SDs)— °C | 38.2±0.71 | 37.7±0.58 | 0.011 |
| Distribution of peak temperature — no./total no. (%) | .. | .. | 0.046* |
| <37.3°C | 4 (7.1%) | 22 (23.9%) | .. |
| 37.3–38.0°C | 21(37.5%) | 54 (58.7%) | .. |
| 38.1–39.0°C | 24(42.9%) | 16 (17.4%) | .. |
| >39.0°C | 7 (12.5%) | 0 | .. |
| Symptoms at onset — no. (%) |  |  |  |
| Cough | 34 (60.7%) | 36 ( 39.1% ) | 0.011 |
| Sputum production | 3 (5.4%) | 11 ( 12.0% ) | 0.183 |
| Nasal congestion | 4 (7.1%) | 2(2.2%) | 0.137 |
| Sore throat | 11(19.6%) | 4(4.4%) | 0.003 |
| Shortness of breath | 10(17.9%) | 4(4.4%) | 0.006 |
| Stomach ache | 1(1.8%) | 2(2.2%) | 0.871 |
| Chest pain | 5 (8.9%) | 3(3.3%) | 0.139 |
| Chest congestion | 10(17.9%) | 13(14.1%) | 0.544 |
| Nausea or vomiting | 4 (7.1%) | 3(3.3%) | 0.281 |
| Diarrhea | 11(19.6%) | 7(7.6%) | 0.03 |
| Myalgia or arthralgia | 17 (30.4%) | 6(6.5%) | <0.0001 |
| poor appetite | 16 (28.6%) | 6(6.5%) | <0.0001 |
| Headache | 2 (3.6%) | 7(7.6%) | 0.319 |
| Fatigue | 32 (57.1%) | 25(27.2%) | <0.0001 |
| Chills | 5 (8.9%) | 5(5.4%) | 0.412 |
| Palmic | 2 (3.6%) | 6(6.5%) | 0.441 |
| Perspire | 1(1.8%) | 0 | 0.198 |
| Cough | 34 (60.7%) | 36 ( 39.1% ) | 0.011 |
| Sputum production | 3 (5.4%) | 11 ( 12.0% ) | 0.183 |
| Comorbidity — no. (%) |  |  |  |
| Any | 24 (42.9%) | 28 (30.4%) | 0.187 |
| Chronic obstructive pulmonary disease | 1 (1.8%) | 2 (2.2%) | 0.871 |
| Diabetes | 4 (7.1%) | 3 (3.3%) | 0.281 |
| Hypertension | 13 (23.2%) | 17 (18.5%) | 0.487 |
| Coronary heart disease | 4 (7.1%) | 3 (3.3%) | 0.281 |
| Cerebrovascular disease | 1 (1.8%) | 3 (3.3%) | 0.591 |
| Chronic renal disease | 1 (1.8%) | 0 | 0.198 |

Data are mean (SDs), median (IQR), n (%), or n/N (%). P values were calculated by Student t test, Mann-Whitney U test, $\chi^2$ test, or Fisher's exact test, as appropriate. \* $\chi^2$ test comparing all subcategories.

The clinical manifestations at onset of those patients have been observed. There were no asymptomatic cases on admission. Besides, 43/56 (76.8%) of patients were with fever on admission, 31/56 (55.4%) of patients were once with fever over 38.0°C, 34/56 (60.7%) with cough, 11/56 (19.6%) with sore throat, 10/56 (17.9%) with shortness of breath, 17/56 (30.4%) with myalgia or arthralgia, 11/56 (19.6%) with diarrhea, 16 (28.6%) with poor appetite, 32 (57.1%) with fatigue.

There were 42.9% (24/56) of patients with comorbidity. The most common comorbidity was hypertension (23.2%), followed by diabetes (4/56, 7.1%), coronary heart diseases (4/56, 7.1%), kidney diseases (1/56, 1.8%), cerebral infarction (1/56, 1.8%) and chronic obstructive pulmonary disease (1/56, 1.8%).

On admission, lymphocytopenia was less common (in 35.7% of the patients), with a mean lymphocyte count of 1.70±0.72×10^9^ per L in this study. Most of the patients showed elevated levels of C-reactive protein (CRP) at ill onset with a median CRP of 10.12 mg per L (IQR 1.33-16.44). The CRP levels of 33.3% patients were over 10 mg/L. All the patients were examined by chest CT 6±3 days after the onset of disease. As shown in the imaging, 24/56 (42.9%) of patients were with bilateral distribution, followed by 22/56 (39.3%) with local distribution, 11/56 (19.6%) with multiple distribution, 7/56 (12.5%) with unilateral distribution. Meanwhile, ground-glass opacity (53/56, 94.6%) was the most common morphological depiction, followed by 10/56 (17.9%) patchy shadowing, 1/56 (1.8%) interstitial abnormalities. (Table 3)

**Table 3:** Radiographic, Laboratory findings, treatments, and clinical outcomes.

|  | Mild or moderate progressed to severe (56) | Match mild or moderate (92) | P value |
| --- | --- | --- | --- |
| **Radiologic findings** |  |  |  |
| Abnormalities on chest CT — no./total no. (%) |  |  |  |
| Local | 22 (39.3%) | 10 (10.9%) | <0.0001 |
| Multiple | 11 (19.6%) | 52 (56.5%) | <0.0001 |
| Bilateral | 24 (42.9%) | 72 (78.3%) | <0.0001 |
| Unilateral | 7 (12.5%) | 18 (19.6%) | 0.374 |
| Patchy shadowing | 10 (17.9%) | 7 (7.6%) | 0.034 |
| Ground-glass opacity | 53 (94.6%) | 82 (89.1%) | 0.432 |
| Interstitial abnormalities | 1 (1.8%) | 19 (20.7%) | 0.002 |
| **Laboratory findings** |  |  |  |
| White-cell count, median (IQR), $\times 10^9$ per L | 5.75(4.66 -7.91) | 5.96 (5.04-7.09) | 0.848 |
| Lymphocyte count, mean (SDs), $\times 10^9$ per L | 1.70±0.72 | 1.88±0.54 | 0.244 |
| Distribution of lymphocyte count— no./total no. (%) | 42 | 90 | 0.468 |
| <1.5$\times 10^9$ per L | 15 (35.7%) | 24 (26.7%) | .. |
| $\geq 1.5 \times 10^9$ per L | 27 (64.3%) | 66 (73.3%) | .. |
| Platelet count, median (IQR), $\times 10^9$ per L | 238 (181-331) | 244 (205-320) | 0.724 |
| Distribution of platelet count— no./total no. (%) | 42 | 90 | 0.183 |
| <150 × 10<sup>9</sup> per L | 2 (4.8%) | 1 (1.1%) | .. |
| ≥150 × 10<sup>9</sup> per L | 40 (95.2%) | 89 (98.9%) | .. |
| Median hemoglobin (IQR) — g per dl | 134 (131-147) | 137 (128-144) | 0.828 |
| C-reactive protein, median (IQR), mg per L | 10.12 (1.33-16.44) | 1.14 (0.37-2.61) | <0.0001 |
| Distribution — no./total no. (%) | 42 | 90 | <0.0001* |
| C-reactive protein ≥10 mg per L | 14 (33.3%) | 5 (5.6%) | .. |
| C-reactive protein <10 mg per L | 28 (66.7%) | 85 (94.4%) | .. |
| **Treatments** | 42 | 90 | .. |
| Antibiotics, antiviral and traditional Chinese medicine | 16 (38.1%) | 40 (50.6%) | 0.188 |
| Antibiotics and antiviral | 5 (11.9%) | 3 (3.9%) | 0.091 |
| Antiviral and traditional Chinese medicine | 6 (14.3%) | 17 (21.8%) | 0.319 |
| **Clinical outcomes at data cutoff (2020.3.12)** |  |  |  |
| — no. (%) |  |  |  |
| Nucleic acid test | 52 | 92 | 0.106 |
| Positive | 3 (5.8%) | 1 (1.1%) | .. |
| Negative | 49 (94.2%) | 91 (98.9%) | .. |
| CT scan | .. | .. | <0.0001* |
| Recovery | 2 (3.8%) | 2 (2.2%) | .. |
| Improved | 48 (92.3%) | 90 (97.8%) | .. |
| Deterioration or new lesions | 2 (3.8%) | 0 | .. |

Data are mean (SDs), median (IQR), n (%), or n/N (%). P values were calculated by Student t test, Mann-Whitney U test, $\chi^2$ test, or Fisher's exact test, as appropriate. \* $\chi^2$ test comparing all subcategories.

According to the symptoms and signs, patients received different treatments. There were 16/42 (38.1%) of patients with antibiotics, antivirus and traditional Chinese medicine, 5/42 (11.9%) of patients with antivirus and antibiotics, 6/42 (14.3%) of patients with antivirus and traditional Chinese medicine. There were 14 patients with no medicine records due to the short time of staying in mobile cabin hospital.

On March 12, those 56 patients were followed up by telephone and 4 lost. There were 5.8% (9/52) of the patients remaining the positive result. As shown in the CT scan, 96.2% (50/52) of the patients were recovery or improved, only 3.8% (2/52) of the patients got deterioration or new lesions. The clinical manifestations and laboratory findings were shown in Table 3.

### The risk factors of patients transforming from mild or moderate type to severe type

The risk factors from univariable analysis with statistic difference were included in multiple regression analysis. Compared with mild or moderate patients, odds of mild-to-severe were higher in patients with cough and fatigue in univariable analysis. Patients of fever on cabin admission, and high level of CRP (≥10 mg per L) were also associated with mild-to-severe type (Table 4). Furthermore, the shelter period of patients less than 7 days was related with high odds transforming severe type. We then included those 148 patients with complete data for top five most statistically significant variables in the multivariable logistic regression model. The results showed that patients with fever on admission of Fangcang shelter hospital, less shelter period (7 days), fatigue and high level of CRP (≥10 mg per L) were associated with increased odds of mild-to-severe type (Table 4).

**Table 4:**
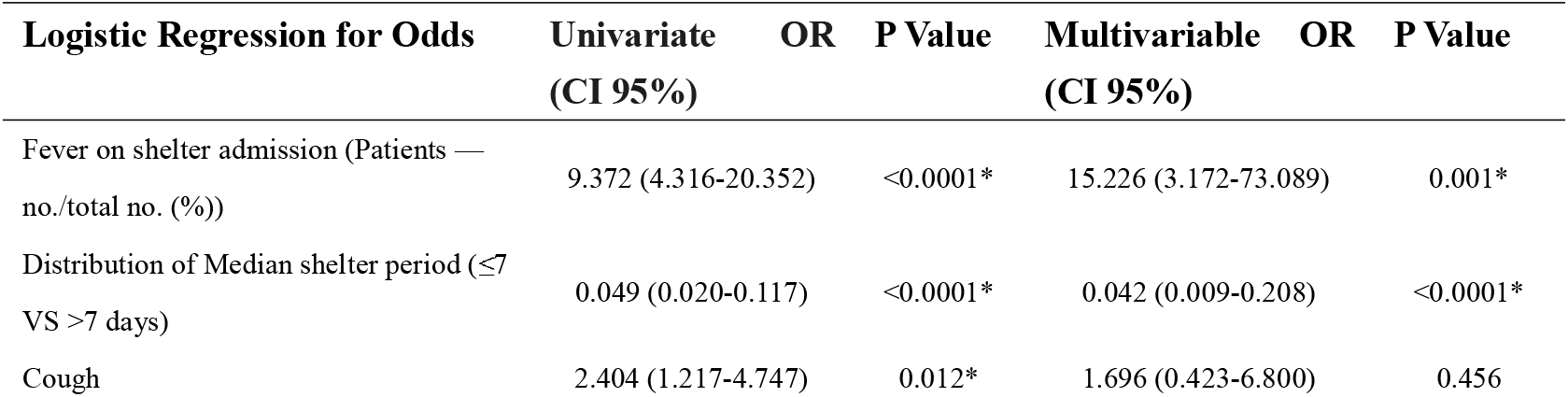
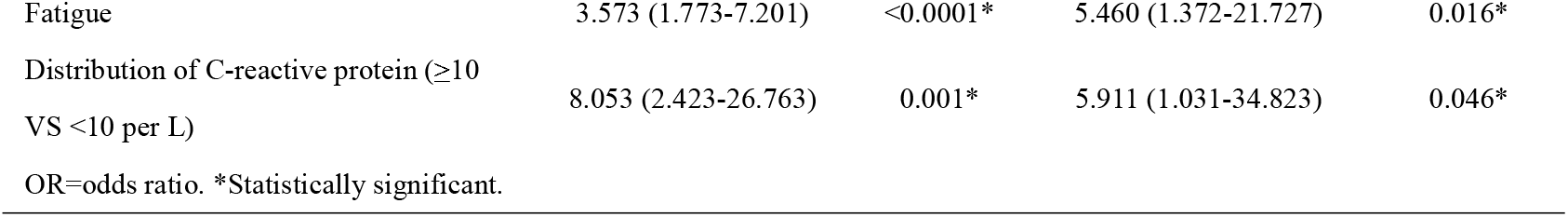
Risk Factors Associated with Mild or moderate pregressed to Severe Transfer from Fangcang Shelter Hospital to the Designated Hospital.

## Discussion

It is the first time to disclose the clinical characteristics and risk factors for developed COVID-19 patients (in Jianghan Fangcang shelter hospital) transferring to the designated hospital. A total of 1,848 COVID-19 patients diagnosed with mild or moderate type were enrolled. After entering the Fangcang shelter hospital, 71.8% (1327/1848) of patients met the criteria for isolation and discharge in the following 35 days. The conversion rate of negative within 14 days was 85.2% among those patients. This effective measure of patient recovery is partly attributed to the characteristics of Jianghan Fangcang shelter hospital. The hospital adopted district management and provided medical service 24 hours on call. The digital ratio of patients to doctors and nurses is about 1.6 to 1. Accurate assessment before admission and effective bed distance in the mobile cabin to some extent prevented the interactions between the patients.^4^ Tens of thousands of new patients have been diagnosed in the world recently every day, which requires enormous medical resources. However, the lack of medical resources in short time is an important cause for the rapid spread and the increased mortality of COVID-19. The Fangcang shelter hospitals could supply the basic treatment for those expanding cases and triage to prevent more family or community transmission cases, in a manner of saving medical resources to the greatest extent.

The novel coronavirus COVID-19 rapidly spread globally, affecting now nearly every continent. The number of patients with severe type determines the final mortality rate of COVID-19. In this study, the case-severity rate was observed with a relatively large prospective cohort, which might be a valuable complement to the characteristics of COVID-19. In the Fangcang shelter hospital of Jianghan, about 3.0% of the patients transformed to severe, which was significantly lower than the 14% cases classified as severe or critical in the spectrum of COVID-19 disease.^8,9^ Furthermore, the median incubation (onset to shelter) period was 10 days and over half of the patients were less than 14 days. Meanwhile, the median time of staying in mobile cabin was 4 days, (21.8%) less than 3 days, 52.7% between 3-7 days, 25.5% more than 7 days. This suggests that the difference in incubation time at the onset of the disease is more indicative for the mild or ordinary patients transforming to severe type. The important function of Fangcang shelter hospital is frequent monitoring and rapid referral. The Jianghan Fangcang shelter hospitals were integrated into the overall health systems of Wuhan via simple pathways of transfer. Overall, Fangcang shelter hospitals had substantially reduced the time from the onset of severe symptoms to admission to a designated hospital, compared to the alternative of home isolation.^10,11^

Previously, older age (over 65 years) was associated with higher odds of progression to severity of COVID-19, which also has been reported as an important independent predictor of mortality in SARS and MERS.^12-14^ In order to better display the clinical characteristics of mild or moderate to severe patients below 65 years old, 92 age-and sex-matched mild or moderate patients stayed in the Fangcang shelter hospital at the same time. In this study, several factors in adults who were hospitalized in Fangcang shelter hospital were associated with mild progressed to severe COVID-19. In particular, patients aged 40-65 years constituted the highest proportion within the severe group in this study. It have reported that 75% of COVID-19 death cases previously suffered 1-2 underlying diseases, a majority of which were diabetes and cardiovascular diseases.^15^ In line with above evidence, our study also found that 42.9% of the mild to severe patients had 1-2 basic diseases, such as cardiovascular diseases, cerebrovascular diseases and endocrine system diseases.

The most common symptom on admission was fever and 76.8% (43/56 cases) mild to severe patients got fever on cabin admission. However, some patients with Covid-19 did not have fever abnormalities on initial presentation, which has complicated the diagnosis.^16^ In this study, 55.4% patients were with peek temperature more than 38.0°C, while 12.5% of patients were once with fever over 39.0°C. High fever was associated with the development of severity and critical death.^17-19^ Therefore, keep vigilance of those mild patients whose peak temperature over 38.0°C. They were quickly transferred to designated higher-level hospitals once the blood oxygen saturation of those patients was less than 93% in Fangcang shelter hospital of Jianghan.

For more specialized monitoring, chest imaging and laboratory services were applied in the Fangcang shelter hospitals. Ground-glass opacity (53/56, 94.6%) was the most common morphological depiction in CT scan on admission. However, only 19.6% of patients showed multiple lesions, 42.9 % patients presented bilateral lesions. Compared with mild patients, most severe patients took CT scan in this study within seven days. The lesions that were present in asymptomatic individuals progressed to bilateral diffuse disease with consolidation around day 10 after the symptom onset.^20-22^ The predominant CT pattern was unilateral and multifocal ground-glass opacities in early stage, then lesions quickly evolved to bilateral, diffuse ground-glass opacity in later stage.^20^ However, those characteristics were not consistent with what we had expected in this study. Therefore, the value of lung CT in determining the prognosis of mild COVID-19 patients still needs further research. Meanwhile, most of the patients had elevated levels of CRP at ill onset with 33.3% over 10 mg/L. Similarly, compared to mild or moderate cases, severe cases more frequently had higher levels of CRP.^16,23-24^ Therefore, imaging and laboratory results could contribute to make the quick decision of transferring to the designated hospitals.

Notably, several clinical manifestations were identified as risk factors for progression from mild to severe in the univariate logistic regression analysis. We found that fatigue on admission was associated with increased odds of mild to severe (Table 4). Furthermore, 57.1% patients had the manifestation fatigue. Report had showed that 45% severe survivors still had cough on discharge and 72% severe non-survivors still had cough at the time of death.^16,17^ In this study, we found cough is associated with case-severity outcome of COVID-19 in univariable analysis. Less common symptoms include a sore throat myalgia or arthralgia, poor appetite. However, respiratory system affection remained as the primary symptom.^23,25-26^ Overall, onset of fever and fatigue symptoms should be closely monitored among cabin hospital, more attention should also be paid to patients on those isolation patients at home. Moreover, the shelter period of patients less than 7 days was related with high odds transforming severe type.

There were some limitations in this study. Firstly, due to limited medical resources in Fangcang shelter hospital, not all laboratory tests have been performed, such as lactate dehydrogenase, IL-6, and serum ferritin. Secondly, the study was limited to the patients with mild or moderate infection in a single center study. However, we focused on the therapeutic and preventive value of mobile cabin hospital in the COVID-19 patients with mild or moderate type. The study population is representative of cases mild developed to severe in Wuhan. To the best of our knowledge, this is the largest retrospective cohort study to disclose the clinical characteristics and risk factors for developed COVID-19 patients transferring to the designated hospital in Fangcang shelter hospital. Overall, Fangcang shelter hospitals have substantially reduced the time from the onset of severe symptoms to admission to a designated hospital. Early application of the Fangcang shelter hospital may contribute to alleviate the shortage of medical resources and decrease the ratio of severe patients. We believe that the information in the article will provide valuable data for other countries facing the COVID-19 pandemic, when they are establishing the national public health emergency management for COVID-19.

## Contributors

HS designed the study and had full access to all of the data in the study and was responsibility for the integrity of the data and the accuracy of the data analysis. HW, JH, YW, ZW, XC, CY and XF collected data. BW, YL and YF analyzed data. YL and YF wrote the Article. All authors critically revised the manuscript for important content and gave final approval for the version to be published.

## Data Availability

The data that support the findings of this study are available from the corresponding author upon reasonable request.

## Acknowledgments

No specific grant from any funding was applied to this research. The Jianghan Fangcang shelter hospital was managed daily by Union Hospital, Tongji Medical College, Huazhong University of Science and Technology and twenty-one medical teams from other provinces associated with six local hospitals participated in the work. We thank 1573 staff in the Jianghan Fangcang shelter hospital, including 308 doctors, 872 nurses, 120 medical technician, 16 hospital infection-control staff and 257 administrative logistical staff.

